# Exploring U.S. food system workers’ intentions to work while ill during the early COVID-19 pandemic: a national survey

**DOI:** 10.1101/2022.04.25.22274276

**Authors:** Caitlin A. Ceryes, Jacqueline Agnew, Andrea L. Wirtz, Daniel J. Barnett, Roni A. Neff

## Abstract

**Background:** While “stay at home” orders were in effect during early phases of the COVID-19 pandemic, many U.S. food workers attended in-person work, charged with maintaining operation of the national food supply chain. Anecdotal evidence suggests that many U.S. food system workers encountered barriers to staying home despite symptomatic COVID-19 illness.

**Methods:** We conducted a national, cross-sectional, online survey between July 31 to October 2, 2020, among 2,535 respondents. We used multivariable regression and free-text analyses to explore factors associated with U.S. food system workers’ intentions to attend work while ill (i.e., presenteeism intentions) during the first four to six months of the COVID-19 pandemic.

**Results:** Overall, 8.8% of workers surveyed reported intentions to attend work while symptomatic with COVID-19 disease. Almost half of respondents (41.1%) reported low or very low household food security. Workers reporting a high workplace safety climate score were half as likely to report presenteeism intentions (adjusted odds ratio [aOR] 0.52, 95% confidence interval (CI) 0.37, 0.75) relative to those reporting low scores. Workers reporting low (aOR 2.06, 95% CI 1.35, 3.13) or very low (aOR 2.31, 95% CI 1.50, 3.13) levels of household food security had twice the odds of reporting presenteeism intentions relative to those reporting high/marginal food security.

**Conclusions:** Our findings suggest that workplace culture and safety climate could enable employees to feel like they can take leave when sick during a pandemic, which is critical to individual health and prevention of workplace disease transmission. However, the pressure experienced by food workers to work when ill, especially by those experiencing food insecurity, themselves, underscores the need for strategies which address these vulnerabilities and empower food workers to make health-protective decisions.

## INTRODUCTION

After the March 11^th^, 2020 World Health Organization COVID-19 pandemic declaration (World Health Organization, 2020), the United States (U.S.) government deemed food system workers, i.e., those responsible for producing, processing, distributing, selling, and serving food, “essential” (1). To maintain operation of the national food supply chain, many U.S. food workers attended work in person while “stay at home orders” were in effect during the early COVID-19 pandemic, Consequently, essential food workers experienced high levels of COVID-19 exposure and illness risks (2), largely due to inability to socially distance while working (3–5). Presenteeism, a phenomenon wherein employees attend work despite symptomatic illness (6), is an important risk factor for workplace and community COVID-19 spread (7,8), especially in workplaces with limited social distancing. Anecdotal evidence suggests that many food system workers encountered barriers to staying home if ill (9). Understanding factors associated with the intent to work while ill (here termed “presenteeism intentions”) is important for preventing workplace spread of COVID-19 and other infectious illnesses.

### U.S. Food System Workforce

In the U.S. food system, approximately 21.5 million workers produce, process, distribute, sell, and serve food in mostly “non-relocatable” jobs (10,11). Appendix 1 provides food sector and subsector characteristics. Despite doing diverse tasks across sectors and jobs, many food workers share demographic and occupational similarities and all are needed to maintain a functioning food supply chain. Studying these workers as a group rather than in occupational silos provides insights relevant to this large cohort, their individual sectors, and food system functionality and resiliency.

Even before the COVID-19 pandemic, many food system workers experienced challenges associated with negative outcomes (12–14), including presenteeism (15). Food system jobs are often characterized by: full-time wages at or below the poverty line (16–18) (Appendix 1); low unionization rates, job insecurity, and at-will employment (19); precarious tipped work (11) or piece work (20); and lacking sick pay and health insurance (21). These jobs exhibit high injury and illness rates relative to national averages (22,23), despite suspected widespread reporting suppression (24,25) and surveillance exemptions (26). Many food jobs exist in the “gig economy,” meaning they are commonly exempted from many labor protections (27) and occupational health surveillance (28).

### Presenteeism

Early presenteeism research examined economic and productivity losses resulting from employees working while sick or injured (29). More recently, studies have investigated the implications of presenteeism for food safety (30) and for worker and community health (6,31). Pre-pandemic studies found that organizational factors (e.g. work policies or cultures), job characteristics (e.g. shift design, job demands), and personal characteristics (e.g. financial stability concerns, personal sense of duty, and perceived co-worker expectations) (15) can potentiate presenteeism.

A limited literature explores presenteeism among food system workers, identifying associations between presenteeism and high work demands; poor employer-employee communication; poor staffing; inadequate workplace policies (e.g., lacking paid sick leave or requiring doctors’ notes) (30,32,33); poor workplace safety climate (34); and job insecurity, job dissatisfaction, and hazardous working conditions (35). During the COVID-19 pandemic, one study of restaurant workers has found that expanding paid sick leave at a large restaurant chain reduced presenteeism when compared to similar chains (36). Other studies among non-healthcare worker cohorts suggest that COVID-19 presenteeism is associated with household income, food security, and age (37), poor access to health benefits (37) and poor workplace safety climate (38). Despite their importance for maintaining national food security, high occupational vulnerability, and concerns about COVID-19 spread, little is known about how food system workers navigated decisions to attend work if ill.

### COVID-19 Presenteeism-Related Policies

At the time of survey, COVID-19 case rates and deaths were rising (39) and the Centers for Disease Control and Prevention (CDC) had issued guidance for sick workers to stay home or isolate (40). However, concerns remained that exacerbated financial pressures and other factors could incentivize presenteeism (41,42). In April 2020, the federal government implemented the first national sick leave policy (43) and augmented unemployment insurance (44), providing paid sick leave for many food chain workers who previously lacked this benefit, including part-time workers (43,45). However, firms employing fewer than 50 or more than 500 people were excluded from this policy, and voluntary implementation was inconsistent (36). Additionally, many processing workers were encouraged or required to work with COVID-19 symptoms (5,46) following a presidential executive order preventing closures of meat and poultry processing plants (47).

To our knowledge, no study has examined presenteeism intentions in a large, nationwide, food system worker sample. Here we explore workplace and non-workplace factors associated with food system worker COVID-19 presenteeism intentions during the early COVID-19 pandemic to identify opportunities to support food workers to remain home if ill or at risk of infecting others.

## METHODS

We drew data from the Johns Hopkins COVID-19 Food Worker Survey, developed during the early COVID-19 pandemic and deployed from July 31, 2020 to October 2, 2020. This cross-sectional, national, online survey of 3,399 food system workers documented COVID-19 pandemic-related workplace experiences and conditions. Recruitment and survey design have been reported in-depth elsewhere (2).

### Study population

The survey included individuals who worked in any of six targeted food system sectors (production; processing; distribution; retail; service; assistance), were literate in English or Spanish, lived in the U.S., were 18 years old or older, and had attended a food system job in-person since March 11th, 2020.

Of 3,831 who initiated the survey, 25.4% of respondents did not answer the question corresponding to presenteeism intentions; thus, their data were excluded from analyses. We also excluded participants who had previously contracted COVID-19, and/or did not receive a paycheck, producing an analytic sample of 2,535 participants. Participants missing outcome data were more likely to identify as Hispanic/Latinx and/or work at organizations with fewer than 10 employees than those with outcome data. Missing outcome data was not associated with age, race, gender, U.S. census region, having worked in the past month, or degree of customer interaction. We discuss missing data patterns for independent variables and implications for interpretations in the discussion.

Sample size calculations determined that a sample of at least 1,000 respondents would provide enough power to detect group differences using a 3% margin of error and 95% confidence for the outcome. The median survey completion time was 19.5 minutes.

### Instrument

In brief, the 114 item instrument was created with input from workers, worker representatives and experts in survey design, disaster preparedness, and occupational health. We used validated scales where possible and included novel items to capture COVID-19-related perspectives about working conditions. Measures are summarized below.

### Measures

Demographics included age, gender identity, race, ethnicity, highest educational attainment, household income, and geographic location. All questions included “don’t know” or “not applicable” options and participants could skip any item beyond demographics. The survey was terminated if demographic responses did not satisfy inclusion criteria. Appendix 2 presents survey items and coding.

### Presenteeism Intentions

We derived our main outcome from the level of agreement with the statement: “If I was sick with COVID-19, but I was still able to work, I would go to work.” The five-point Likert scale was dichotomized to: workers who strongly agreed or agreed with the statement versus all others.

### Occupational Measures

Workers indicated their food system sector and subsector from an edited Food Chain Workers Alliance list (FCWA; a coalition of food worker-based organizations) (11). Workers employed in more than one sector were asked to indicate the job in which they worked the most hours. Occupational characteristics included job tenure, full/part-time status, organization size, customer contact, work transportation, whether workers were told they were “required” to work by their employers, and union membership. Respondents specified all workplace benefits provided by their employers since the pandemic declaration from a select-all-that-apply list (48); these were aggregated as frequencies and analyzed individually.

We assessed quantitative work demands and workplace social support using medium-length scales from the Copenhagen Psychosocial Questionnaire III (COPSOQ III) (49), following published scoring procedures and then dichotomizing scores at the median into “high” and “low” categories. Higher work demands scores indicated more challenging levels of work demands (e.g., time pressure or many overlapping tasks). We assessed organizational safety climate using a 6-item scale (50) where high scores indicated that workers perceived their organization had a high commitment to safety. We created a composite organizational safety climate variable by summing scale responses and dichotomizing at the median, including responses for all participants who had answered 5 or more (of six) items.

### Non-Occupational Measures

We measured food security since March 11, 2020, using a United States Department of Agriculture (USDA) Six-item Short Form Household Food Security Survey Module (51). The composite categorical variable included responses of participants with two or more items (of six) and was scored according to USDA classifications: high/marginal food security (raw score 0-1); low food security (2 – 4); and very low food security (5 – 6). Cronbach’s alpha was > 0.7 for all scales except quantitative work demands, which was 0.67 (52).

We measured attitudes regarding reopening the economy based on agreement with the statement, “It is worth the health risk to reopen the economy as soon as possible.” The 6-point Likert scale was collapsed to three points: agreement; neither agreement nor disagreement; and disagreement.

### Theoretical Approach

The Job Demands-Resources (JD-R) (53) and Total Worker Health (TWH) models (54,55) guided analyses. The JD-R model suggests that job resources can mitigate the negative health impacts of workplace demands (53). We therefore hypothesized that resources such as organizational safety climate (defined as employees’ shared perceptions of their organization’s prioritization of worker safety (50,56) and paid sick leave would reduce the likelihood of reporting presenteeism intentions. The Total Worker Health approach (55) considers external (non-workplace) factors that impact worker well-being. Our conceptual model (presented in Ceryes et al., 2021 (2)) includes workplace and non-workplace factors associated with food worker outcomes, including presenteeism, during the COVID-19 pandemic.

### Statistical analyses

We used STATA 14 I/C (College Station, Texas USA) for quantitative analyses. Statistics included Chi^2^ or Rank Sum tests (significance value p < 0.05) as well as Spearman’s rank and Pearson’s correlation coefficients to identify collinearity and bivariate logistic regression to assess correlations according to presenteeism intention status. Adjusted logistic regression models were used to examine associations with workplace characteristics. Variables associated with the outcome, presenteeism intention, at the level of P<.05 were retained in the multivariable model (age, gender, food system sector, organization size, hourly status).

Additional covariate inclusion was informed by *a priori* conceptual associations (race, ethnicity, geographic location). We included food security status and perspectives on reopening the economy based on free-text data (described below) and bivariable associations (P <0.05). The final model estimated associations between presenteeism intentions and workplace, and non-workplace characteristics while controlling for age, race, ethnicity, gender, food system sector, organization size, and hourly status. Akaike’s Information Criteria (AIC) values were used to assess model fit, and variance inflation factors assessing multicollinearity were all less than four (mean = 1.43) (57).

Sensitivity analyses were conducted by stratifying on degree of customer interaction and whether workers were told they were “required” to work and controlled for clustering at the state level. Estimates did not meaningfully differ from our primary results (Appendix 3).

### Free-text Analyses

Many survey participants provided detailed responses to the open-ended question: “*Do you have any other comments about the level of risk from COVID-19, or decisions about whether to go to work?*” These comments often included discussion of presenteeism intentions; thus, we analyzed responses to elaborate on our quantitative findings (58), an approach used previously in survey-based presenteeism studies (59). Comments informed covariate selection by narrowing variables considered for analyses. For example, responses frequently mentioned food insecurity and perspectives on opening the economy; therefore, we retained those variables. We also used comments to choose between highly correlated variables (e.g., food security status over annual household income), and when interpreting and discussing quantitative results.

The lead investigator (CAC) conducted two close reviews of free-text data, taking notes before coding responses and organizing them into themes (60) and excluding non-substantive comments (e.g. “N/A” or “No”). Atlas.ti (Version 8.0, Berlin, Germany) and Microsoft Excel (Washington, USA) were used to sort, organize, and manage free-text data. Respondents offering comments were compared to those who did not and to the full sample to identify potential biases. We analyzed presenteeism-related text responses as a whole and by sector, by subgroups according to reports of presenteeism, and by benefits and working conditions. Qualitative memos tracked CAC’s reactions to comments (61).

The Johns Hopkins Bloomberg School of Public Health Institutional Review Board considered this study exempt (category 2) (IRB No. 12549).

## RESULTS

### Quantitative Results

Table 1 presents analytic sample demographics. Respondents were primarily female (64.8%), not Hispanic/Latinx (90.0%), white (86.0%), non-union (79.6%), working full-time (64.8%) and of average age 45.9 years (SD 11.2). Most worked in restaurant/service (43.3%) and retail (34.9%), with the fewest in distribution (2.4%). Almost all (95.9%) had worked in-person in the past month before taking the survey. Nearly a third (32.7%) were told they were “required” to work by their employers at some point between pandemic onset and the survey in August-September, 2020. Almost half of respondents (41.1%) reported low or very low food security. Analytic sample demographics resembled those of the overall study population.

**Table 1.**
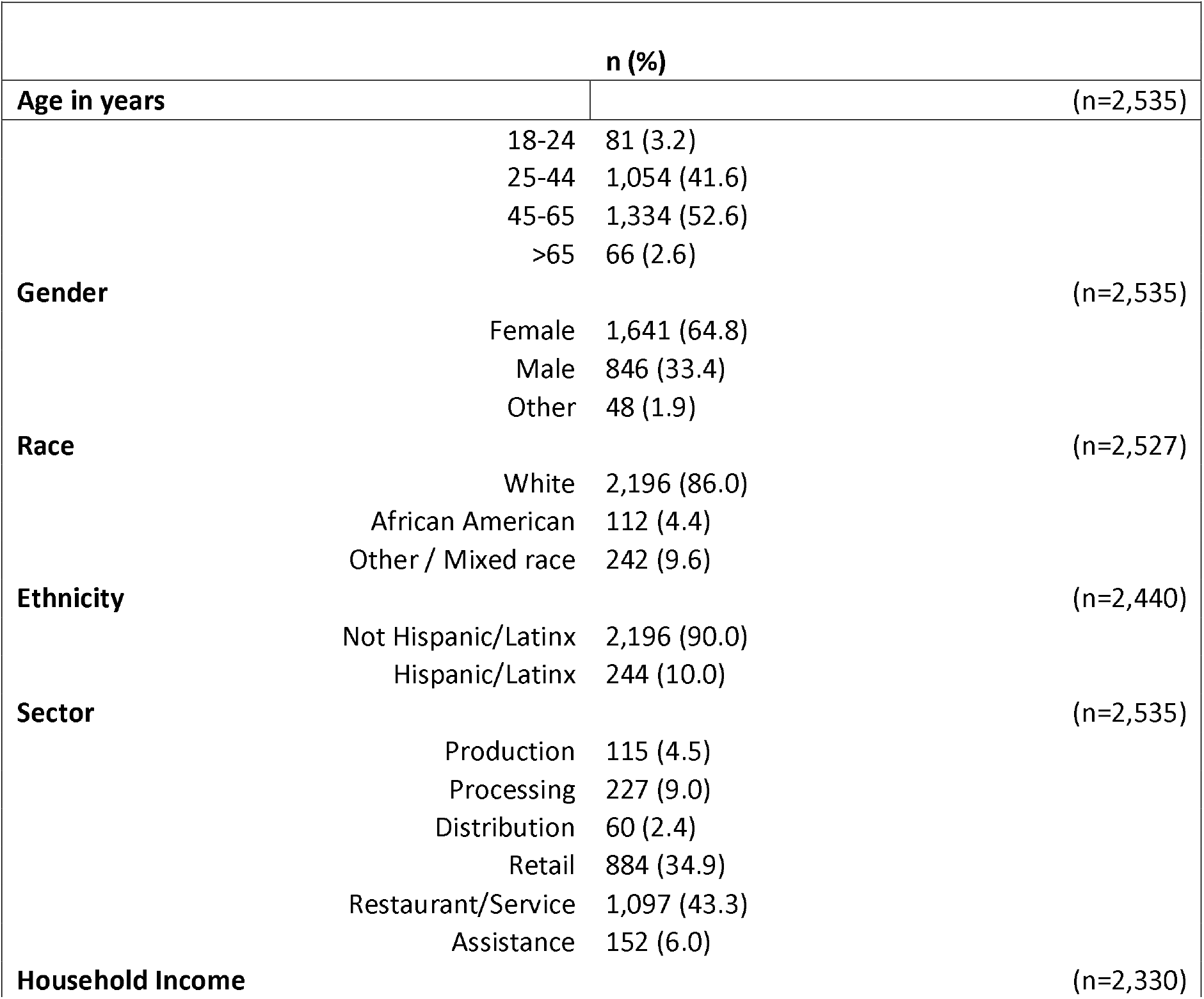
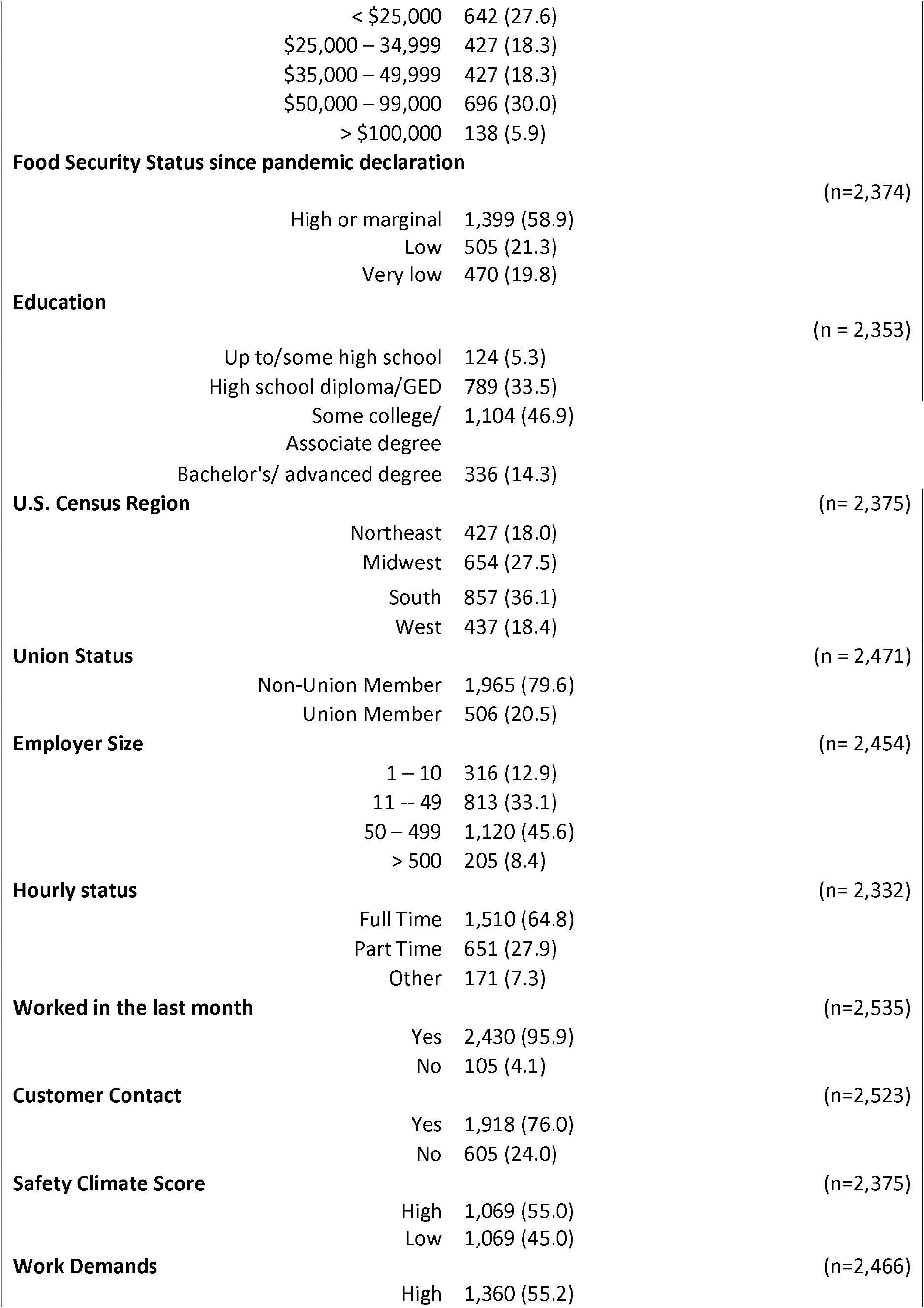
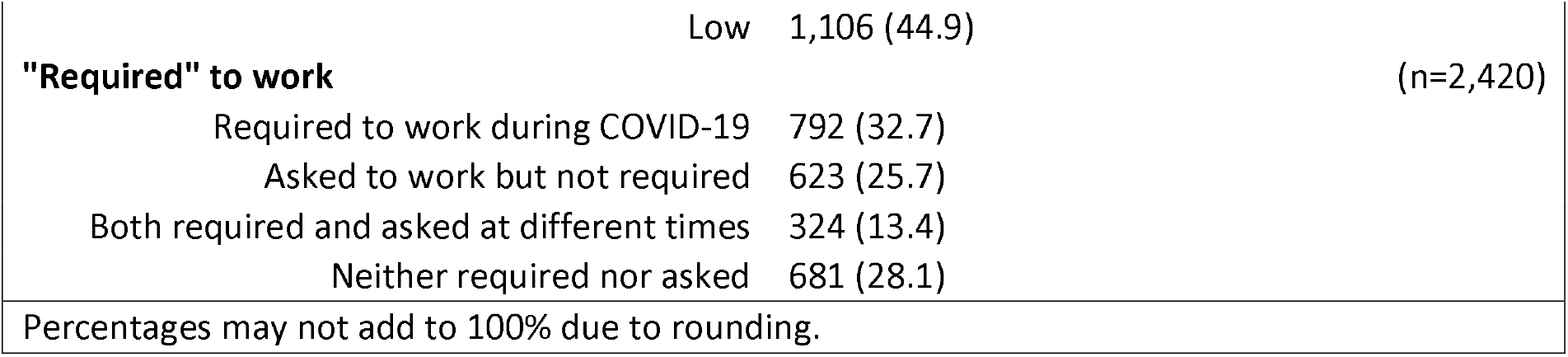
Demographic and occupational characteristics for a national U.S. food system worker cohort during early COVID-19.

|  |  |  |
| --- | --- | --- |
|  | **n (%)** |  |
| **Age in years** |  | (n=2,535) |
|  | 18-24 | 81 (3.2) |
|  | 25-44 | 1,054 (41.6) |
|  | 45-65 | 1,334 (52.6) |
|  | >65 | 66 (2.6) |
| **Gender** |  | (n=2,535) |
|  | Female | 1,641 (64.8) |
|  | Male | 846 (33.4) |
|  | Other | 48 (1.9) |
| **Race** |  | (n=2,527) |
|  | White | 2,196 (86.0) |
|  | African American | 112 (4.4) |
|  | Other / Mixed race | 242 (9.6) |
| **Ethnicity** |  | (n=2,440) |
|  | Not Hispanic/Latinx | 2,196 (90.0) |
|  | Hispanic/Latinx | 244 (10.0) |
| **Sector** |  | (n=2,535) |
|  | Production | 115 (4.5) |
|  | Processing | 227 (9.0) |
|  | Distribution | 60 (2.4) |
|  | Retail | 884 (34.9) |
|  | Restaurant/Service | 1,097 (43.3) |
|  | Assistance | 152 (6.0) |
| **Household Income** |  | (n=2,330) |

|  |  |  |  |
| --- | --- | --- | --- |
| | < \$25,000 | 642 (27.6) | |
| | \$25,000 – 34,999 | 427 (18.3) | |
| | \$35,000 – 49,999 | 427 (18.3) | |
| | \$50,000 – 99,000 | 696 (30.0) | |
| | > \$100,000 | 138 (5.9) | |
| **Food Security Status since pandemic declaration** |  |  | (n=2,374) |
|  | High or marginal | 1,399 (58.9) |  |
|  | Low | 505 (21.3) |  |
|  | Very low | 470 (19.8) |  |
| **Education** |  |  | (n = 2,353) |
|  | Up to/some high school | 124 (5.3) |  |
|  | High school diploma/GED | 789 (33.5) |  |
|  | Some college/<br>Associate degree | 1,104 (46.9) |  |
|  | Bachelor's/ advanced degree | 336 (14.3) |  |
| **U.S. Census Region** |  |  | (n= 2,375) |
|  | Northeast | 427 (18.0) |  |
|  | Midwest | 654 (27.5) |  |
|  | South | 857 (36.1) |  |
|  | West | 437 (18.4) |  |
| **Union Status** |  |  | (n = 2,471) |
|  | Non-Union Member | 1,965 (79.6) |  |
|  | Union Member | 506 (20.5) |  |
| **Employer Size** |  |  | (n= 2,454) |
|  | 1 – 10 | 316 (12.9) |  |
|  | 11 -- 49 | 813 (33.1) |  |
|  | 50 – 499 | 1,120 (45.6) |  |
|  | > 500 | 205 (8.4) |  |
| **Hourly status** |  |  | (n= 2,332) |
|  | Full Time | 1,510 (64.8) |  |
|  | Part Time | 651 (27.9) |  |
|  | Other | 171 (7.3) |  |
| **Worked in the last month** |  |  | (n=2,535) |
|  | Yes | 2,430 (95.9) |  |
|  | No | 105 (4.1) |  |
| **Customer Contact** |  |  | (n=2,523) |
|  | Yes | 1,918 (76.0) |  |
|  | No | 605 (24.0) |  |
| **Safety Climate Score** |  |  | (n=2,375) |
|  | High | 1,069 (55.0) |  |
|  | Low | 1,069 (45.0) |  |
| **Work Demands** |  |  | (n=2,466) |
|  | High | 1,360 (55.2) |  |
|  | Low | 1,106 (44.9) |  |
| **"Required" to work** |  |  | (n=2,420) |
| Required to work during COVID-19 |  | 792 (32.7) |  |
| Asked to work but not required |  | 623 (25.7) |  |
| Both required and asked at different times |  | 324 (13.4) |  |
| Neither required nor asked |  | 681 (28.1) |  |
| Percentages may not add to 100% due to rounding. |  |  |  |

### Presenteeism

Of 2,535 respondents, 8.8% agreed that they would attend work if sick with COVID-19, but these differed greatly by sector. The production sector had the highest proportion of workers reporting presenteeism intentions (24.2%), followed by processing workers (10.6%), restaurant and service workers (8.3%), and retail workers (7.5%). Food assistance workers were least likely to report presenteeism intentions (3.7%), (x^2^=45.31, p<0.001). Appendix 4 shows comparisons between groups reporting presenteeism intentions versus not, by variables of interest.

### Benefits

Of 2,527 respondents, 27.7% reported paid sick leave access, 30.1% reported “easier” access to sick leave since March 11th, 2020, and 14.0% had received free workplace COVID-19 testing since the pandemic declaration.

### Multivariable Model

Table 2 presents bivariate (Model 1) and multivariable logistic regression (Models 2 and 3) results for variables of interest (organizational safety climate; work demands; access to paid leave; food security; perspectives about reopening the economy) and presenteeism intentions, adjusted for age, gender, ethnicity, race, full/part-time status, food system sector and organization size. See Appendix 5 for all models.

**Table 2.**
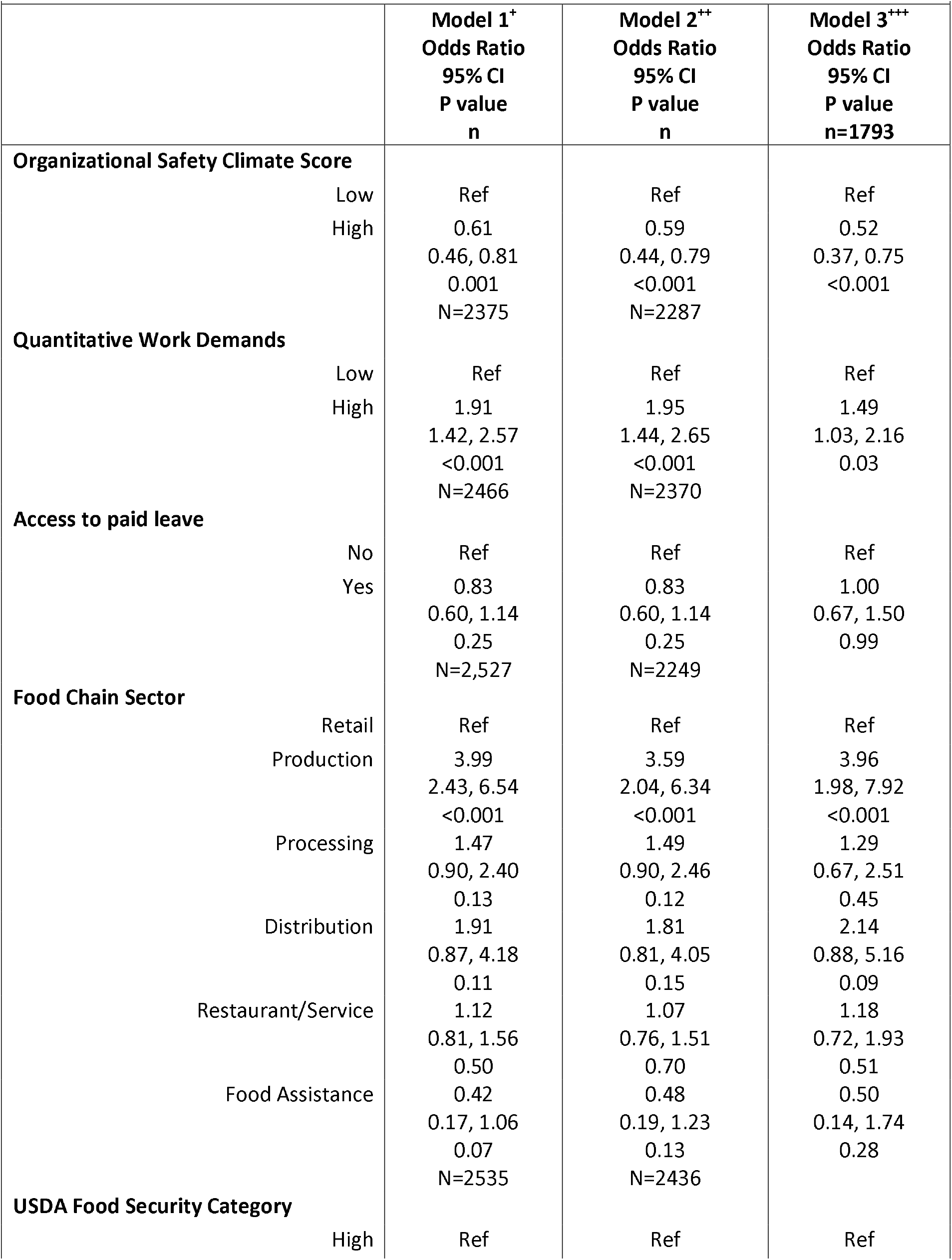
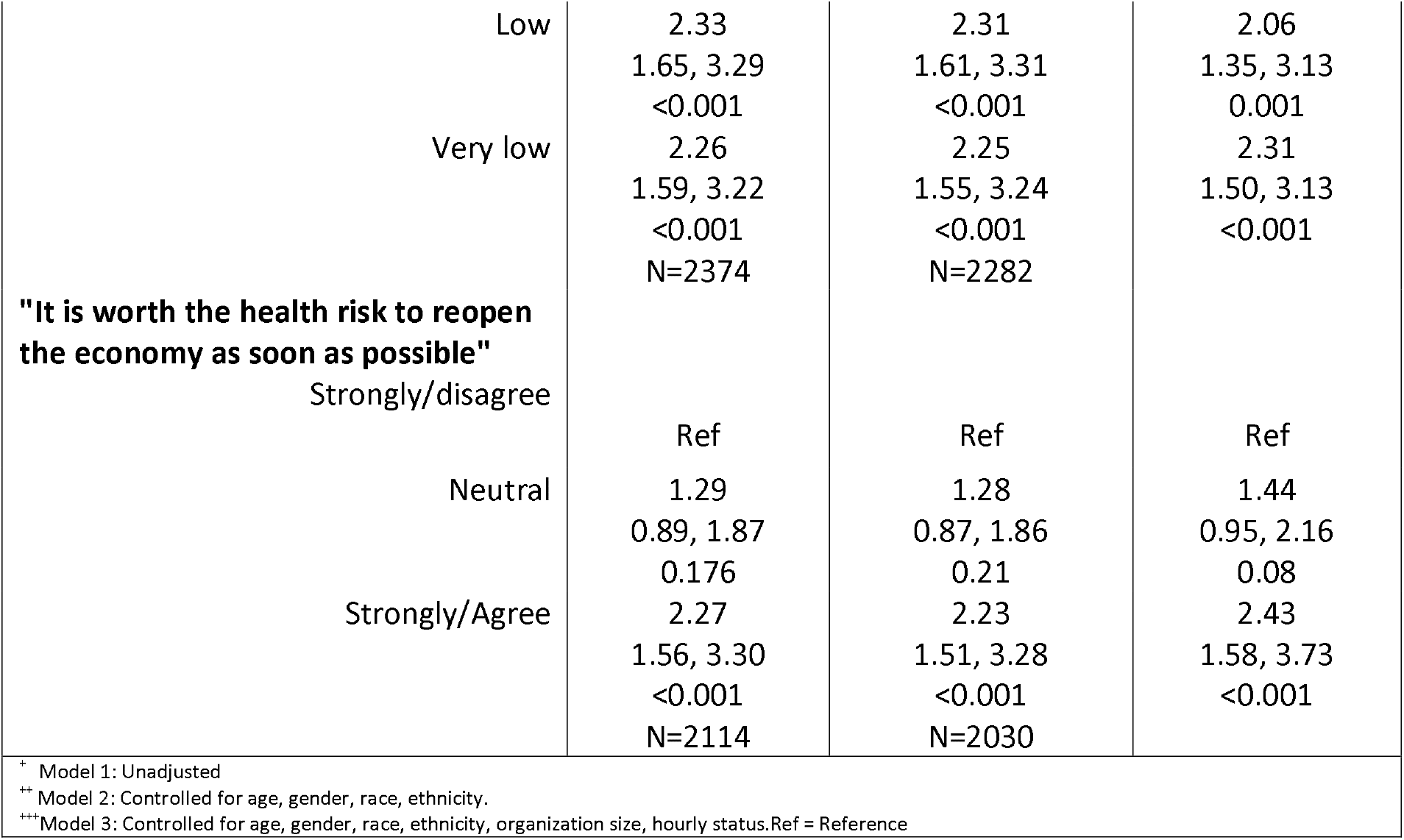
Workplace and non-workplace factors associated with reporting presenteeism intentions in a national food chain worker sample during early COVID-19.

After adjustment, respondents reporting high levels of organizational safety climate were half as likely to report presenteeism intentions compared to those reporting lower scores (adjusted odds ratio [aOR] 0.52, 95% CI 0.37, 0.75). Workers with high levels of work demands had 49% greater odds of reporting presenteeism intentions relative to those reporting lower levels (aOR 1.49, 95% 1.03, 2.16). Food production workers had higher odds of reporting presenteeism intentions relative to retail workers (aOR 3.96; 95% CI 1.98, 7.92). Paid sick leave was not associated with presenteeism intentions.

Respondents reporting low or very low food security were more than twice as likely to report presenteeism intentions relative to those reporting marginal/high food security (aORs 2.06, 95% CI 1.35, 3.13 and 2.31, 95% CI 1.50, 3.13, respectively). Respondents agreeing or strongly agreeing that it was “worth the health risk” to reopen the economy had higher odds of reporting presenteeism intentions relative to those disagreeing with this statement (aOR 2.43, 95% CI 1.58, 3.73).

### Free Text Results

Overall, 13.5% answered the question, “*Do you have any other comments about the level of risk from COVID-19, or decisions about whether to go to work?*” with 460 substantive comments. Responses ranged from 1 to 233 words, with 23-word median length. Production workers had the lowest word count (13 words) and retail the highest (24 words). Workers who commented were less likely to work in food production, report annual household incomes below $15,000 or above $100,000, and were more likely to work for tips and report very low food security status (Appendix 6.)

### Workplace factors

Policies: Many comments mentioned employers’ policies relating to presenteeism and workplace COVID-19 spread. While a few workers described adequate sick pay if symptomatic or COVID-19-positive, others described insufficient policies and benefits, including lacking paid sick leave. A retail worker explained, “*Obviously no one wants to go to work sick, but it is necessary since the pay is so low and I don’t get sick pay*.” Respondents also described barriers to quarantine and testing. For example, a retail worker described financial disincentives to disclosing exposure, “*If I was to be exposed to someone with Covid I would not tell my [employers] about it because they will not pay me to be off work. I cannot afford to be off work*.” Barriers to testing included unpaid isolation periods, *“*… *it is a 2 week or more wait for results. If you are tested you may not return to work until you get results. How many people with mild symptoms are going to be out of work for 2 weeks or more voluntarily?”* (restaurant worker), and high test costs, *“The test cost as much as half of my weekly wage”* (retail worker). Others described policies providing only partial sick pay, or requiring employees to find shift coverage, use personal vacation time, obtain doctors’ notes, or abide penalty-driven attendance systems. For example, a processing worker described: “*If you were sick or had any of the symptoms of COVID-19, if you didn*’*t go to work they would “point” [penalize] you for that so if you have enough points you will eventually* ‘*point out’ [lose your job]*.”

Culture: Even if employers had official policies supporting those who stayed home, employees described cultural factors which communicated an expectation to work when symptomatic. For example, workers cited concerns about employer retaliation for using sick leave: “*Calling in sick is frowned upon. People who call in sick frequently get less hours [meaning less pay] and the worse [less desirable] hours”* (retail worker). A restaurant worker described related job insecurity, *“Even if you don*’*t get fired for calling out … they’ll find something else to fire you for…* “

Some comments described instances where policies meant to discourage COVID-19 presenteeism were unclear or not followed, including symptomatic co-workers working after symptom-checks, and managers ignoring COVID-19 symptoms rather than sending staff home. One retail worker’s superior explicitly discouraged testing, “*Boss told us not to get tested so we wouldn’t have to miss work*.”

### Non-workplace factors

#### Economic precarity

Aside from workplace conditions, workers cited economic instability, stemming from insufficient wages, as a driver for presenteeism. Many comments mentioned the need to make ends meet, working paycheck to paycheck, and working to buy food for their families. A processing worker explained, “*There is NO decision!*… *We have bills and children to feed…I cannot stay home!*” A retail worker shared her frustration about having used up her sick leave, meaning she had limited options, “*what the **** am I gonna do, not feed my kids?*… *(pardon my profanity, it’s necessary for emphasis, I can’t really convey how strongly I feel about this*.)”

Distrust of public health messaging: Some respondents viewed COVID-19 as exaggerated or not a credible health threat. As one processing worker described, “*I think it’s blown out of proportion and has very skewed and inaccurate testing. I don’t think I’m anymore at risk than the seasonal flu*.”

## DISCUSSION

Our findings identify workplace and non-workplace conditions associated with food system workers’ intentions to work while ill and provide insights into this decision. While our results are specific to the COVID-19 pandemic context, we believe they have relevance for both infectious disease outbreak planning and mitigating the spread of more quotidian contagions.

Given rapid changes in infection rates, resources available for worker protection, and scientific knowledge about COVID-19 throughout 2020 and 2021, it is important to view these results in their temporal context. This study occurred during the first four to six months of the pandemic, when vaccines were unavailable, after initiation of federal paid sick leave policies, and before eviction moratoriums and unemployment insurance enhancements expired (62). Because of rapid U.S. case rate increases and news coverage emphasizing disease severity during these months (63), respondents may have perceived COVID-19 as more severe than other illnesses and planned to remain home. As the pandemic continued and many states prioritized “reopening,” essential and non-essential workers were encouraged to return to work, and supporting policies were relaxed or rescinded. Therefore, if repeated later in the pandemic, a similar study might show an even greater prevalence of presenteeism intentions among these workers.

## Workplace Factors associated with Presenteeism Intentions

### Organizational Safety Climate

Workers with high safety climate scores, indicating they perceived their employers valued and prioritized maintaining employees’ safety, were less likely to report COVID-19 presenteeism intentions. This finding aligns with other pre- and mid-pandemic studies suggesting that organizational safety climate influences workers’ presenteeism decisions (38,64,65) and builds on previously established connections between safety climate and COVID-19 safety perceptions (2).

Free-text data elaborated on how organizational safety climate constructs, here including employees’ shared perceptions of safety priorities, policies, and procedures; managerial commitment to safety; employee behavioral norms, and worker safety activity participation (50), could influence presenteeism intentions. For example, comments describing managers who ignored COVID-19 safety policies indicate a lack of empowerment to participate in safety activities and policy enforcement. Organizational safety climate is often studied regarding its effects on injury prevention, but these findings suggest its underlying constructs could represent important intervention targets for reducing illness-related presenteeism.

### Sick Leave

The lack of association between sick leave access and presenteeism intentions after adjustment was surprising. Workers’ comments describing cultural and organizational barriers to using sick leave, even if it is “officially” established, provide one interpretation of this finding. Descriptions of retaliation and penalties barring workers from accessing sick leave indicate that some employees are not empowered to use it. Such barriers have been documented among restaurant workers (45), and we expand these findings to include other food system workers. Our results diverge from those of Schneider and colleagues’ (2021), who found that increasing paid sick leave reduced COVID-19 presenteeism among restaurant workers at the Olive Garden fast-casual restaurant chain. We suggest the difference could again relate to empowerment because Olive Garden’s paid sick leave expansion occurred following “significant public scrutiny,” (36), their employees might have felt more able to access their newfound benefits than workers whose employers were not being scrutinized.

### Work Requirements

Notably, 32.7% of respondents reported being told they were “required” to work during the COVID-19 pandemic. Because these workers lacked choice, this circumstance would not typically be considered presenteeism. Sensitivity analysis estimates of reported presenteeism, stratified by requirement to work, did not meaningfully differ from our primary results. Research should assess the physical and mental health impacts of requirements to work during the COVID-19 pandemic.

### Sector differences

After controlling for demographics and job characteristics, production workers were more likely to report presenteeism intentions relative to retail workers. This finding could relate to reduced risk perceptions due to these workers’ open-air working environments and not interacting with customers. Alternatively, H-2A visa holders (meaning those in the United States on temporary agricultural work visas) might feel obliged to attend work while ill in order to remain in the country (66). Research is needed to explore this association further. We did not identify other sector-specific differences or note differential comment content by sector, though production workers were less likely to provide comments than workers in other sectors.

### External factors associated with presenteeism intentions

#### Food Security

Over 40% of respondents reported experiencing low or very low food security, despite working at in-person food jobs during the COVID-19 pandemic. After controlling for covariates, these workers were more than twice as likely to report presenteeism intentions than those with marginal or high food security. This finding, combined with many free-text comments that mentioned the need to work to buy food, suggests food insecurity was a major driver of presenteeism intentions in this population. Our findings align with Tilchin and colleagues’ (2021) findings that perceived food insecurity was associated with a three-fold increase in intention to work sick among U.S. employees. The paradox of food workers experiencing food insecurity while feeding the nation has been previously acknowledged in literature on farmworkers (67), and we re-emphasize its inherent inequity here. We also note that these findings could help explain broader disparities in COVID-19 morbidity and mortality (68) during early pandemic waves.

### Risk Perceptions

Workers who felt it was “worth the health risk” to reopen the economy were twice as likely to report presenteeism intentions. Comments suggested some respondents did not trust public health messaging about COVID-19’s severity, and/or felt the benefits of working, including financial stability, outweighed COVID-19 exposure risks. This finding highlights connections between effective and consistent public health messaging and reducing infectious disease spread.

### Future research and recommendations

Longitudinal studies should further examine the association between workplace culture and presenteeism, especially whether shifts in workplace safety climate can decrease the spread of workplace and community infectious disease. Research is also needed to explore ways to empower employees to fully participate in developing and enacting policies, such as paid sick leave and symptom checks, especially in the context of top-down federal or state policy mandates and prolonged emergencies or pandemics.

This study suggests that worker food insecurity represents a major driver of COVID-19 presenteeism intentions. We therefore endorse instituting and evaluating policies that improve workers’ overall financial stability to prevent presenteeism and accompanying disease transmission. These policies include raising food workers’ compensation to a living wage, limiting “just in time” shifts and standardizing work schedules so that workers can plan for childcare and other needs, and providing reliable, full-time, benefitted work to those who want it (69). Such actions would not only contribute to public health and food system stability but could also reduce food businesses’ presenteeism-related economic losses, which are estimated to be substantial (29). Finally, we advocate for heightened external accountability around workplace safety protocols and practices, including proactive worksite inspections and statutory worker protections, especially for “essential” workplaces. It would be informative to track presenteeism and its associated influences and outcomes in a longitudinal manner should a similar national disaster occur in the future.

While this large national survey addresses the experience of a unique worker population that is critical to our food supply, there are some expected limitations. As with many other Internet-based surveys, our sample overrepresented white, female, and high-income individuals (70,71). Despite efforts to minimize missing data, thus increasing sample size and diversity, few participants identified as African American and Hispanic/Latinx or other Black/Indigenous/People of Color (BIPOC) individuals. These groups are of great interest because they are believed to be more subject to the negative impacts of COVID-19 (72). This study may have underestimated levels of risk factors or the existence of presenteeism intentions, especially among these populations. Future studies must focus on including these groups.

Use of free text data always presents the challenge of interpretation, especially when a single coder reviews the responses. However, our text analyses related directly to our validated scales and served the purpose of expanding, clarifying, and prioritizing those results.

This cross-sectional study was conducted during the early stages of the pandemic, when COVID-19 knowledge and risk perception were evolving, and anxiety was high. Although the design does not allow for causal inferences, results during this critical period indicate participants’ perceptions of causal relationships between several risk factors and presenteeism decisions.

## CONCLUSIONS

The COVID-19 pandemic has highlighted U.S. society’s reliance on food system workers to maintain national food security. Despite their heightened risks for COVID-19 morbidity and mortality, many food system workers indicated they would attend work while ill during the early COVID-19 pandemic. Often, they felt that they had no choice. This research suggests that interventions targeting workplace safety climate and food insecurity among food system workers could reduce presenteeism, therefore protecting the national food supply and the public’s health during the COVID-19 pandemic and in other disaster or infectious illness scenarios. Addressing barriers to staying home when ill, like improving safety climate and mitigating or eliminating vulnerabilities such as food insecurity, could enable food system workers to make decisions that protect both themselves and their workplaces. Reducing presenteeism is critical for creating optimal worker health outcomes, public health outcomes, and maintaining a functioning food system.

## Data Availability

All data produced in the present study are available upon reasonable request to the authors

## Declarations

### Consent for Publication

Not applicable.

### Availability of Data and Materials

The datasets used and/or analysed during the current study are available from the corresponding author on reasonable request.

### Authors’ Contributions

Conceived study, obtained funding: RAN. Designed the study: RAN, CAC, DJB. Data collection: CAC, RAN. Led data analysis and manuscript writing: CAC. Contributed to analysis: RAN, ALW, JA. Participated in writing and revisions: RAN, AJW, JA, DJB.

## Acknowledgments

We thank the researchers at the Labor Occupational Health Program at the University of California, Berkeley, Celeste Monforton from George Washington University, and Jeffrey Johnson for their survey development contributions. We thank United Commercial and Food Worker Union and Food Chain Workers Alliance representatives, Martha Ojeda, and other food workers for providing survey input, and Bytes.co for survey dissemination collaboration. Thank you to Dr. Steven Harvey for free-text analysis guidance, and to Joelle Robinson and Erin Biehl for their contributions to study activities including recruitment, coding, and incentive administration.

