## Appendix 1. U.S. Food System Workforce for "Exploring U.S. food system workers’ intentions to work while ill during the early COVID-19 pandemic: a national survey" (from another version of this preprint: version 2, 2022-05-09)

### Supplemental Materials

**Appendix 1. U.S. Food System Workforce:** The U.S. food system is an interconnected system which relies on approximately 21.5 million workers who produce, process, distribute, sell, and serve food in mostly "non-relocatable" jobs ^3,4^. Table A1 provides an overview of food system sector and subsector characteristics. When considered together, these workers form one of the nation's largest employment segments ^4^. Despite engaging in diverse work tasks across sectors and jobs, many individuals engaged in food system work share various demographic and occupational similarities. For this reason, studying these workers as a group rather than in occupational silos may provide insights relevant to this large worker cohort, the individual sectors that comprise it, and the functionality and resilience of the food system itself.

| **Table A1.** Food system sectors, wage and employment estimates, and main sub-sectors included in the survey with North American Industrial Classification System (NAICS) codes. | | | | |
| --- | --- | --- | --- | --- |
| Sector | 2015 Annual and Hourly Wages (USD) | 2015 Estimated Employment^1^ | Sub-Sectors | NAICS Codes |
| Production | $18,657 annual median;^,^ $11 hourly median^1^ | 2.5 million^1^ | Crop production; animal production; fishing, hunting, trapping | 111, 112, 114 |
| Food Processing | $28,000 annual median; $13 hourly median^1^ | 1.8 million^1^ | Animal food manufacturing; grain and oilseed milling; Sugar and confectionary processing; Fruit and vegetable preserving/specialty food manufacturing; Dairy product manufacturing; Animal slaughtering and processing; Seafood product preparation; Bakeries and tortilla manufacturing; Other food manufacturing | 311 |
| Food Distribution^+^ | $35,000 annual median; $14 hourly median^1^ | 3.3 million^1^ | Trucking; warehousing and storage; wholesale grocery; delivering restaurant food/groceries to homes/businesses | 4244, 4245, 492210 |
| Food Retail | $15,000 annual median; $10 hourly median^1^ | 3.1 million^1^ | Supermarkets, grocery stores, warehouse clubs, supercenters that sell food, convenience stores, and specialty food stores. | 4451, 4452, 4523, 311811 |
| Restaurant/  Service | $12,000 annual median; $9.30 hourly median^1^ | 11 million^1^ | Restaurants (including bar and fast food); catering; institutional food. | 772 |
| Food Assistance | $30444 annual mean^2^ | 33,850^3^  Note: sector is heavily reliant on volunteer labor. | K-12 school food programs; food banks and pantries; food benefits programs; meal delivery or in-person meal provision program. | 624200 |
| ^1^ Because NAICS categories do not overlap fully with whether someone would be considered a food system worker, Food Chain Worker Alliance generated employment estimates for each sector using data from the American Community Survey (2010-2014), Current Population Survey’s Annual Social and Economic Supplement (2003-2016), Outgoing Rotation Groups (2003-2016), and Food Security Supplement (2001-2014), and Occupational Employment Statistics (2015). We consider these 2016 estimates the most comprehensive estimates currently available. Estimation methods can be found in the Food Chain Workers Alliance report, *No Piece of the Pie* (2016) available from <http://foodchainworkers.org/wp-content/uploads/2011/05/FCWA_NoPieceOfThePie_P.pdf>.  ^2^ Author calculation averaging 2020 national wage estimates for 23 occupational titles which include the word "food" in the "Community Food and Housing, and Emergency and Other Relief Services" NAICS code (624200) downloaded from <https://www.bls.gov/oes/current/naics4_624200.htm> on 15 Nov, 2021.  ^3^ Author calculation summing employment estimates for 23 occupational titles which include the word "food" in the "Community Food and Housing, and Emergency and Other Relief Services" NAICS code (624200) downloaded from <https://www.bls.gov/oes/current/naics4_624200.htm> on 15 Nov, 2021. | | | | |
