## Appendix 2. Survey questions used in a study of presenteeism intentions among a large national sample of food chain workers during the first 4-6 month for "Exploring U.S. food system workers’ intentions to work while ill during the early COVID-19 pandemic: a national survey" (from another version of this preprint: version 2, 2022-05-09)

| Appendix 2. Survey questions used in a study of presenteeism intentions among a large national sample of food chain workers during the first 4-6 months of the COVID-19 pandemic. | | | |
| --- | --- | --- | --- |
| **Variable** | **Question** | **Answers** | **Transformation** |
| Presenteeism Intentions | If I was sick with COVID-19, but I was still able to work, I would go to work | 1 = Strongly Agree, 2 = Agree, 3 = Neutral, 4 = Disagree, 5 = Strongly Disagree, 99 = DK | Binary: 0 = Agreed with statement, 1 = Did not agree (includes neutral) |
| **Demographics** | | | |
| Age^+^ | What is your age? | Integer | Categorical: 0 = 18 - 24 years, 1 = 25 - 44 years, 2 = 45 - 64 years, 3 = ≥ 65 years |
| Gender*^+^ | Which of the following best describes your gender identity?  (Select all that apply) | Male, Female, Transgender, Non-binary, Prefer to self-describe | Categorical: 0 = Female, 1 = Male, 2 = Other response |
| Race*^+^ | What is your race? Check all that apply: | White, Black/African American, Asian American, Native American/American Indian or Alaska Native, Native Hawaiian or other Pacific Islander, Other | Categorical: 0 = White, 1 = African American, 2 = Other response/multiple races |
| Ethnicity^+^ | What is your ethnicity? | 1 = Yes, Hispanic, 0 = No, Not Hispanic | NA |
| Income* | Which of the following best describes your household income range in 2019 before taxes? | Less than $10,000, $10,000 to $14,999, $15,000 to $24,999, $25,000 to $34,999, $35,000 to $49,999, $50,000 to $74,999, $75,000 to $99,999, $100,000 to $149,999, $150,000 to $199,999, $200,000 or more | 1 = < $25,000; 2 = $25,000 - $34,999; 3 = $35,000 - $49,999; 4 = $50,000 - 99,999; 5 = ≥100,000 |
| Geographic Location | Census region derived from U.S. zip code or U.S. state | Zip code or U.S. state | Categorical: 1 = Northeast, 2 = South, 3 = Midwest, 4 = West |
| Worked during the past month | Have you worked in the food industry at any point in the past MONTH? | 0 = no; 1 = yes | NA |
| **Occupational Attributes** | | | |
| Job tenure | How many years have you worked at this job? | 1 = Less than one year, 2 = 1-2 years, 3 = 3-5 years, 4 = 6-10 years, 5 = More than 10 years, 99 = Don't know | NA |
| Full/Part Time Status | How would you describe your employment at your food job? | 1 = Full-time, 2 = Part-time, 3 = Other | NA |
| Employer Size* | About how many employees are at your place of work? | 1 = 1 – 10, 2 = 11 – 49, 3 = 50 – 499, 4 = 500 – 999, 5 = More than 1000, 99 = Don't know | Categorical: 1 = 1 - 10; 2 = 11 - 49; 3 = 50 - 499, 4 = ≥500 |
| Food System Sector* | Which part of the food industry is your job in? | 1 = food production, 2 = food processing, 3 = food distribution, 4 = food retail, 5 = restaurant and food service, 6 = food assistance programs, 7 = other, 8 = none | Categorical: 1 = Small/Specialty Store, 2 = Supermarket/Box store |
| Union Membership | Are you represented by a union? | 1 = Yes, 0 = No, 99 = Don't Know | NA |
| Quantitative Work Demands | First, we would like to know amount the amount of work and how busy you have been **since COVID-19.** In your job… | 100 = Always, 75 = Often, 50 = Sometimes, Seldom, 25 = Never/hardly ever, 99 =Don't Know | Continuous: Scores for 3 items calculated according to the COPSOQ III. Guidelines and Questionnaire. |
|  | …how often do you not have time to complete all your work tasks? |  |  |
|  | …do you get behind with your work? |  | Binary: Scores dichotomized at the median value. |
|  | …is your workload unevenly distributed so it piles up? |  |  |
| Organizational Safety Climate | Please rate how much you agree with these statements about health and safety at your work **in the last month.** | 4 = Strongly Agree, 3 = Agree, 2 = Disagree, 1 = Strongly Disagree, 88 = Not Applicable | Continuous: Scores for 6 items averaged according to Hahn & Murphy, 2008. |
|  | New employees learn quickly that they are expected to follow good health and safety practices. |  |  |
|  | Employees are told when they do not follow good safety practices. |  | Binary: Scores dichotomized at the median value. |
|  | Workers and management work together to ensure the safest possible conditions. |  |  |
|  | There are no major shortcuts taken when worker health and safety are at stake. |  | Missing data correction: respondents with ≥5 items considered complete |
|  | The health and safety of workers is a high priority with management where I work. |  |  |
|  | I feel free to report safety problems where I work. |  |  |
| Workplace Social Support | Please indicate how often these things happen at your job, **in the last month**. | 100 = Always, 75 = Often, 50 = Sometimes, Seldom, 25 = Never/hardly ever, 99 =Don't Know | Continuous: Scores for each 2 item scale (manager, coworker, community) averaged and combined according to COPSOQ III. Guidelines and Questionnaire. |
|  | How often do you get help and support from your manager, if needed? |  |  |
|  | How often is your manager willing to listen to your problems at work, if needed? |  | Binary: Scores dichotomized at the median value. |
|  | How often do you get help and support from your co-workers, if needed? |  |  |
|  | How often are your co-workers willing to listen to your problems at work, if needed? |  |  |
|  | Is there a good atmosphere between you and your colleagues? |  |  |
|  | Do you feel part of a community at your place of work? |  |  |
| Customer Interaction | Do you work directly with customers? | 1 = Yes, 0 = No, 99 = I don't know | NA |
| Work transportation | In the past month, which did you use to get to your food job most days? (select the best answer) | 1 = Bus or public transit, 2 = Traveled myself: Drive, walk, or bike, 3 = Carpool, someone drove me, taxi or rideshare app | NA |
| Requirement to work | Did your employer tell you either of these things about coming to work during COVID-19? [check only one] | 1 = Said you are required to come to work during COVID-19, 2 = Asked you to come to work during COVID-19 but did not require it, 3 = Both, at different times, 4 = Neither, 99 = Don't Know | Binary: 0 = too few/about right, 1 = too many |
| **Non-occupational attributes** | | | |
| USDA Food Security Score | The next questions are about the food situation in your household during the COVID-19 pandemic. How true are these statements about your household’s food situation since the COVID-19 outbreak on March 11th? |  | Responses of “often” or “sometimes” on questions 1 &2, and “yes” on 3, 5, 6, are coded as affirmative (yes). Responses of “almost every month” and “some months but not every month” on 4 are coded as affirmative (yes).   The sum of affirmative responses to the six questions in the module is the household’s raw score on the scale. Food security status is assigned as follows:  Raw score 0-1—High or marginal food security (raw score 1 may be considered marginal food security, but a large proportion of households that would be measured as having marginal food security using the household or adult scale will have raw score zero on the six-item scale)  Raw score 2-4—Low food security  Raw score 5-6—Very low food security |
|  | 1. The food that my household bought just didn't last, and I/we didn't have money to get more | 1 = Never true, 2 = Sometimes true, 3 = Often true, 99 = I don't know |  |
|  | 2. I/we couldn't afford to eat balanced meals | 1 = Never true, 2 = Sometimes true, 3 = Often true, 99 = I don't know |  |
|  | 3. Did you (or other adults in your household) ever cut the size of your meals or skip meals because there wasn't enough money for food? (If yes, please indicate how often below.) | 1 = Yes, 0 = No, 99 = I don't know |  |
|  | 4. Since you responded "yes" to cutting the size of your meals since COVID-19, how often did this happen? | 1 = Once, 2 = Twice, 3 = Weekly, 4 = Daily |  |
|  | Since COVID-19: |  |  |
|  | 5. Did you ever eat less than you felt you should because there wasn't enough money for food? | 1 = Yes, 0 = No, 99 = I don't know |  |
|  | 6. Were you ever hungry but didn't eat because there wasn't enough money for food? | 1 = Yes, 0 = No, 99 = I don't know |  |
| Perspectives on opening the economy | It is worth the health risk to reopen the economy as soon as possible. | 1 = strongly disagree, 6 = strongly agree (slider) | Condensed to 1 = Disagree; 2 = Neutral 3 = Agree |
| ^+^Forced response question. |  |  |  |
| *Responses for some questions collapsed due to small sample sizes. |  |  |  |
| Respondents could skip any non-forced question without selecting a "refuse" option. | |  |  |
