## Appendix 3. Sensitivity analyses for associations between variables of interest and odds ratio for COVID-19 presenteeism intentions within a national for "Exploring U.S. food system workers’ intentions to work while ill during the early COVID-19 pandemic: a national survey" (from another version of this preprint: version 2, 2022-05-09)

| **Appendix 3. Sensitivity analyses for associations between variables of interest and odds ratio for COVID-19 presenteeism intentions within a national sample of food system workers in the U.S.** | | | |
| --- | --- | --- | --- |
| **Subgroup:** | **Respondents reporting customer interaction** | **Respondents who were "Required" to work** | **Accounting for state-level clustering** |
|  | **Odds Ratio**  **95% CI**  **P value**  **N=1171** | **Odds Ratio**  **95% CI**  **P value**  **N=664** | **Odds Ratio**  **95% CI**  **P value** |
| **Organizational Safety Climate Score** |  |  |  |
| Low | Ref | Ref | Ref |
| High | 0.70  0.43, 1.11  0.13 | 0.53  0.31, 0.90  0.02 | 0.63  0.42, 0.94  0.02 |
| **Quantitative Work Demands** |  |  |  |
| Low | Ref | Ref | Ref |
| High | 1.52  0.93, 2.51  0.10 | 1.29  1.73, 2.27  0.38 | 1.51  1.00, 2.30  0.05 |
| **USDA Food Security Category** |  |  |  |
| High | Ref | Ref | Ref |
| Low | 1.52  0.85, 2.76  0.16 | 1.26  0.66, 2.41  0.48 | 2.18  1.35, 3.50  0.001 |
| Very low | 2.87  1.62, 5.08  <0.001 | 1.39  0.72, 2.71  0.33 | 2.37  1.41, 3.98  0.001 |
| **Access to paid leave** |  |  |  |
| No | Ref | Ref | Ref |
| Yes | 1.22  0.71, 2.08  0.71 | 0.82  0.44, 1.53  0.53 | 1.18  0.74, 1.87  0.49 |
| **"It is worth the health risk to reopen the economy as soon as possible"**  Strongly/disagree | Ref | Ref | Ref |
| Neutral | 1.45  0.85, 2.49  0.18 | 1.99  1.07, 3.69  0.03 | 1.47  0.93, 2.32  0.10 |
| Strongly/Agree | 2.57  1.46, 4.52  0.001 | 2.63  1.37, 5.05  0.004 | 2.40  1.47, 3.92  <0.001 |
| **Food System Sector** |  |  |  |
| Retail | Ref | Ref | Ref |
| Production | 3.66  1.03, 13.04  0.05 | 2.50  0.69, 9.13  0.16 | 3.73  1.72, 8.11  0.001 |
| Processing | 0.73  0.15, 3.56  0.70 | 0.80  0.31, 2.08  0.65 | 1.19  0.54, 2.62  0.67 |
| Distribution | 2.10  0.66, 6.68  0.21 | 1.97  0.56, 6.94  0.29 | 2.31  0.85, 6.26  0.10 |
| Restaurant/Service | 1.03  0.56, 1.90  0.92 | 0.86  .40, 1.86  0.70 | 1.25  0.72, 2.17  0.43 |
| Food Assistance | 0.51  0.12, 2.41  0.40 | 0.40  .07, 2.21  0.30 | 0.43  0.09, 1.99  0.28 |
