## Appendix 4. Comparisons between groups reporting presenteeism intentions versus not and variables of interest in a national food system worker sample for "Exploring U.S. food system workers’ intentions to work while ill during the early COVID-19 pandemic: a national survey" (from another version of this preprint: version 2, 2022-05-09)

| **Appendix 4.** Comparisons between groups reporting presenteeism intentions versus not and variables of interest in a national sample of food system workers during early COVID-19. | | | | |
| --- | --- | --- | --- | --- |
|  |  | "If I was sick with COVID-19, but I was still able to work, I would go to work." | |  |
|  |  | Did not agree | Agree |  |
|  |  | N=2313 | N= 222 |  |
|  |  | n (%) or Mean (SD) | n (%) or Mean (SD) | p value |
| **Demographics** |  |  |  |  |
| Age, m(sd) |  | 46.06 (11.27) | 43.75 (10.78) | 0.004 |
| Race (n=2527) | White | 1,991 (86.3) | 182 (82.3) |  |
|  | African American | 100 (4.3) | 12 (5.4) |  |
|  | Other / Mixed race | 215 (9.3) | 27 (12.2) | 0.26 |
| Ethnicity (n=2440) | Not Hispanic/Latinx | 2008 (90.3) | 188 (87.4) |  |
|  | Hispanic/Latinx | 217 (9.8) | 27 (12.6) | 0.19 |
| Gender (n=2535) | Female | 1,521 (65.8) | 120 (54.1) |  |
|  | Male | 754 (32.6) | 92 (41.4) |  |
|  | Other Answer | 38 (1.6) | 10 (4.5) | <0.001 |
| Sector (n=2535 | Production | 87 (3.8) | 28 (12.6) |  |
|  | Processing | 203 (8.8) | 24 (10.8) |  |
|  | Distribution | 52 (2.3) | 8 (3.6) |  |
|  | Retail | 818 (35.4) | 66 (29.7) |  |
|  | Restaurant/Service | 1006 (43.5) | 91 (41.0) |  |
|  | Food Assistance | 147 (6.4) | 5 (2.3) | <0.001 |
| Income (n=2330) | < $25,000 | 586 (27.5) | 56 (27.9) |  |
|  | $25,000 – 34,999 | 385 (18.1) | 42 (20.9) |  |
|  | $35,000 – 49,999 | 382 (17.9) | 45 (22.4) |  |
|  | $50,000 – 99,000 | 45 (30.3) | 51 (25.4) |  |
|  | > $100,000 | 131 (6.2) | 7 (3.5) | 0.17 |
| U.S. Census Region (n=2375) | Northeast | 388 (17.9) | 39 (19.1) |  |
|  | Midwest | 599 (27.6) | 55 (27.0) |  |
|  | South | 785 (36.2) | 72 (35.3) |  |
|  | West | 399 (18.4) | 38 (18.6) | 0.97 |
| **Occupational Attributes** |  |  |  |  |
| Union Status (n=2471) | Non-Union Member | 1796 (79.6) | 169 (79.0) |  |
|  | Union Member | 461 (20.4) | 45 (21.0) | 0.84 |
| Employer Size (n=2454) | 1 – 10 | 288 (12.9) | 28 (13.0) |  |
|  | 11 - 49 | 745 (33.3) | 68 (31.6) |  |
|  | 50 - 499 | 1030 (46.0) | 90 (41.9) |  |
|  | More than 500 | 176 (7.9) | 29 (13.5) | 0.04 |
| Full-time/part-time status(n=2332) | Full Time | 1376 (64.5) | 134 (67.3) |  |
|  | Part Time | 609 (28.6) | 42 (21.1) |  |
|  | Other | 148 (6.9) | 23 (11.6) | 0.01 |
| Customer Contact (n=2523) | No | 546 (23.7) | 59 (26.9) |  |
|  | Yes | 1758 (76.3) | 160 (73.1) | 0.29 |
| Access to Paid Leave (n=2527) | No | 1660 (72.0) | 168 (75.7) |  |
|  | Yes | 645 (28.0) | 54 (24.3) | 0.25 |
| Employer granted "easier" access to sick leave (n=2527) | No | 1596 (69.2) | 170 (76.6) |  |
|  | Yes | 709 (30.8) | 52 (23.4) | 0.02 |
| Work Demands (n=2464) |  | 46.0 (24.4) | 54.0 (25.7) |  |
|  |  |  |  | <0.001 |
| Safety Climate (n=2373) |  | 17.7 (4.4) | 15.9 (4.9) | <0.001 |
| Social Support (n=2287) |  | 67.2 (21.8) | 59.4 (23.3) |  |
|  |  |  |  | <0.001 |
| Required to Work (n=2420) | Required | 702 (31.2) | 90 (42.3) |  |
|  | Asked | 580 (26.3) | 43 (20.2) |  |
|  | Both | 292 (13.2) | 32 (15.0) |  |
|  | Neither | 633 (28.7) | 48 (22.5) | 0.006 |
| **Non-occupational attributes** | | | | |
| Food Security Status since pandemic declaration (n=2374) | High or marginal | 1317 (60.7) | 82 (40.2) |  |
|  | Low | 441 (20.3) | 64 (31.4) |  |
|  | Very low | 412 (19.0) | 58 (28.4) | <0.001 |
| It is worth the health risk to reopen the economy as soon as possible. (n=2114) | Strongly/Disagree | 829 (43.0) | 58 (31.4) |  |
|  | Neither Agree/Disagree | 697 (36.1) | 63 (34.1) |  |
|  | Strongly/Agree | 403 (20.9) | 64 (34.6) | <0.001 |
