## Appendix 5. Model results for non-exposure variables considered for and included in the multivariable model. for "Exploring U.S. food system workers’ intentions to work while ill during the early COVID-19 pandemic: a national survey" (from another version of this preprint: version 2, 2022-05-09)

| **Appendix 5.** Model results for non-exposure variables considered for and included in the multivariable model^+^. | | | |
| --- | --- | --- | --- |
|  |  | Presenteeism Intentions  (Measured by agreeing with the statement,  "*If I was sick with COVID-19, but I was still able to work, I would go to work"*) | |
|  |  | Unadjusted Odds Ratio  95% Confidence Interval  P value  N | Adjusted Odds Ratio^+^  95% Confidence Interval  P value  N=1793 |
| Age |  | 0.98  0.97, 0.99  0.004 | 1.00  0.98, 1.01  0.84 |
| Gender |  |  |  |
|  | Female | Ref | Ref |
|  | Male | 1.55  1.16, 2.06  0.003 | 1.19  0.83, 1.70  0.35 |
|  | Other Response | 3.33  1.62, 6.86  0.001  N=2535 | 0.99  0.12, 7.95  0.99 |
| Race |  |  |  |
|  | White | Ref | Ref |
|  | African American | 1.29  0.68, 2.46  0.432 | 1.28  0.60, 2.73  0.52 |
|  | Other or > 1 Race | 1.37  0.91, 2.10  0.134  N=2527 | 0.80  0.41, 1.55  0.50 |
| Ethnicity |  |  |  |
|  | Non-Hispanic/Latinx | Ref |  |
|  | Hispanic/Latinx | 1.32  0.87, 2.04  0.192  N=2440 | 0.78  0.40, 1.52  0.47 |
| Region | Northeast | Ref | -- |
|  | South | 0.91  0.59, 1.40  0.680 | -- |
|  | Midwest | 0.91  0.61, 1.37  0.660 | -- |
|  | West | 0.95  0.59, 1.51  0.821  N=2375 | -- |
| Full/Part-time status |  |  |  |
|  | Full-time | Ref | Ref |
|  | Part-time | 0.71  0.49, 1.01  0.06 | 0.77  0.50, 1.18  0.23 |
|  | Other | 1.60  0.99, 2.56  0.053  N=2332 | 1.41  0.80, 2.50  0.24 |
| Job tenure | Less than one year | Ref | -- |
|  | 1-2 years | 1.51  0.85, 2.68  0.162 | -- |
|  | 3-5 years | 1.75  1.01, 3.05  0.046 | -- |
|  | 6-10 years | 1.46  0.81, 2.65  0.210 | -- |
|  | More than 10 years | 0.97  0.81, 2.65  0.925  N=2334 | -- |
| Household Income | < $25,000 | Ref | -- |
|  | $25,000 – 34,999 | 1.14  0.75, 1.74  0.537 | -- |
|  | $35,000 – 49,999 | 1.24  0.82, 1.86  0.99 | -- |
|  | $50,000 – 99,000 | 0.83  0.56, 1.23  0.348 | -- |
|  | ≥ $100,000 | 0.56  0.25, 1.25  0.159  N=2330 | -- |
| Organization Size |  |  |  |
|  | 10 or fewer employees | Ref | Ref |
|  | 11 -- 49 | 0.94  0.59,1.49  0.788 | 0.84  0.46, 1.53  0.57 |
|  | 50 – 499 | 0.9  0.58, 1.40  0.637 | 0.87  0.47, 1.62  0.66 |
|  | More than 500 | 1.69  0.98, 2.94  0.061  N=2454 | 1.14  0.49, 1.65  0.77 |
| Sector | Retail | Ref | Ref |
|  | Production | 3.99  2.43, 6.54  <0.001 | 3.96  1.98, 7.92  <0.001 |
|  | Processing | 1.47  0.90, 2.40  0.13 | 1.29  0.67, 2.51  0.45 |
|  | Distribution | 1.91  0.87, 4.18  0.11 | 2.14  0.88, 5.16  0.09 |
|  | Restaurant/Service | 1.12  0.81, 1.56  0.50 | 1.18  0.72, 1.93  0.51 |
|  | Food Assistance | 0.42  0.17, 1.06  0.07  N=2535 | 0.50  0.14, 1.74  0.28 |
| Customer Contact | No | Ref | -- |
|  | Yes | 0.84  0.62, 1.15  0.283 | -- |
| Quantitative Work Demands Score   Low | | Ref | Ref |
|  | High | 1.91  1.42, 2.57  <0.001  N=2466 | 1.49  1.03, 2.16  0.03 |
| **Workplace Safety Climate Score**   Low | | Ref | Ref |
|  | High | 0.61  0.46, 0.81  0.001  N=2375 | 0.52  0.37, 0.75  <0.001 |
| Social Support Score*  Low | | Ref |  |
| High | | 0.98  0.98, 0.99  <0.001  N=2289 | -- |
| Workplace Benefits since March 11 | | N=2,111 |  |
|  | Extra “hazard pay” separate from overtime pay | 0.92  0.70, 1.21  0.550 | -- |
|  | Access to paid sick leave | 0.76  0.55, 1.04  0.086 | 1.00  0.67, 1.50  0.99 |
|  | Made it easier to take sick leave if needed | 0.66  0.48, 0.91  0.010 | -- |
|  | Paid family sick leave | 0.95  0.59, 1.52  0.827 | -- |
|  | Free/discounted food | 1.26  0.89, 1.81  0.213 | -- |
|  | Health Insurance | 0.96  0.66, 1.39  0.832 | -- |
|  | Free COVID-19 Testing | 1.25  0.86, 1.80  0.242 | -- |
|  | Covering medical costs if an employee gets COVID-19 | 0.92  0.57, 1.49  0.746 | -- |
|  | Child/Elder Care | 1.46  0.67, 3.14  0.334 | -- |
|  | Private Transportation | 1.39  0.49, 3.99  0.538 | -- |
| **"It is worth the health risk to reopen the economy as soon as possible"** | Strongly/disagree | Ref | Ref |
|  | Neutral | 1.29  0.89, 1.87  0.176 | 1.44  0.95, 2.16  0.08 |
|  | Strongly/Agree | 2.27  1.56, 3.30  <0.001  N=2114 | 2.43  1.58, 3.73  <0.001 |
| Transportation | Bus or public transit | Ref | -- |
|  | Drive/walk/bike (traveled self) | 0.62  0.28, 1.40  0.255 | -- |
|  | Carpool/taxi/rideshare | 0.72  0.27, 1.91  0.507 | -- |
| ^+^Fully adjusted model includes: Workplace Safety Climate, work demands, access to paid sick leave, food security score, perspectives on reopening the economy, age, gender, race, ethnicity, food system sector, hourly status, and organization size.  Ref = Reference value | | | |
