## Appendix 6. Comparison of participants who self-selected to provide additional comments on their decision to attend work. for "Exploring U.S. food system workers’ intentions to work while ill during the early COVID-19 pandemic: a national survey" (from another version of this preprint: version 2, 2022-05-09)

| Appendix 6. Comparison of participants who self-selected to provide additional comments on their decision to attend work with non-commentors and entire sample. | | | | | |
| --- | --- | --- | --- | --- | --- |
|  |  | Commentors | Non-commentors |  | Total Sample |
|  |  | n (%) or mean, SD | n (%) or mean, SD | p value | n (%) or mean, SD |
| Total N |  | N=460 | N=2,939 |  | N=3,399 |
| Sector | | | | | |
| (n=460) | Production | 15 (3.3) | 155 (5.3) |  | 170 (5.0) |
|  | Processing | 39 (8.5) | 262 (8.9) |  | 301 (8.9) |
|  | Distribution | 21 (4.6) | 61 (2.1) |  | 82 (2.4) |
|  | Retail | 155 (33.7) | 1000 (34.0) |  | 1,155 (33.4) |
|  | Restaurant/  Service | 200 (43.5) | 1285 (43.7) |  | 1,485 (43.7) |
|  | Assistance | 30 (6.5) | 176 (6.0) | 0.02 | 206 (6.1) |
| Age |  |  |  |  |  |
| (n=460) | Mean, SD | 44.5 (11.2) | 46.1 (11.3) | 0.003 | 46.0 (11.3) |
| Gender | | | | | |
| (n=460) | Female | 317 (68.9) | 1890 (64.3) |  | 2,207 (65.0) |
|  | Male | 134 (29.1) | 985 (33.5) |  | 1,119 (32.9) |
|  | Other | 9 (2.0) | 64 (2.2) | 0.156 | 73 (2.2) |
| Race | | | | | |
| (n=459) | White | 386 (84.1) | 2504 (85.6) |  | 2,890 (85.4) |
|  | Non-White | 73 (15.9) | 426 (14.6) | 0.525 | 495 (14.6) |
| Ethnicity |  |  |  |  |  |
| (n=448) | Hispanic | 43 (9.6) | 2514 (89.0) |  | 353 (10.8) |
|  | Non-Hispanic | 405 (90.4) | 310 (11.0) | 0.382 | 2,919 (89.2) |
| Income | | | | | |
| (n=445) | <10,000 | 10 (2.2) | 93 (4.8) |  | 103 (4.3) |
|  | 10,000 - 15,000 | 27 (6.1) | 150 (7.7) |  | 177 (7.4) |
|  | 15,-000 - 24,999 | 71 (16.0) | 310 (15.9) |  | 381 (15.9) |
|  | 25,000 - 34,999 | 88 (19.8) | 353 (18.1) |  | 441 (18.5) |
|  | 35,000 - 49,999 | 78 (17.5) | 356 (18.3) |  | 434 (18.2) |
|  | 50,000 - 74,999 | 97 (21.8) | 385 (19.8) |  | 482 (20.2) |
|  | 75,000 - 99,999 | 56 (12.6) | 173 (8.9) |  | 229 (9.6) |
|  | >= 100,000 | 18 (4.0) | 125 (6.4) | 0.02 | 143 (6.0) |
| U.S. Region | | | | | |
| (n=443) | Northeast | 80 (18.1) | 488 (17.9) |  | 568 (17.9) |
|  | South | 119 (26.9) | 758 (27.8) |  | 877 (27.7) |
|  | Midwest | 170 (38.4) | 975 (35.7) |  | 1145 (36.1) |
|  | West | 74 (16.7) | 507 (18.6) | 0.661 | 581 (18.3) |
| Union Membership | | | | | |
| (n=451) | Non-union | 354 (78.5) | 2285 (80.1) |  | 2639 (79.6) |
|  | Union | 97 (21.5) | 567 (19.9) | 0.423 | 664 (20.5) |
| Job Pay Status | | | | | |
| (n=441) | Hourly | 325 (73.7) | 1468 (75.4) |  | 1793 (75.1) |
|  | Hourly with tips | 49 (11.1) | 49 (2.5) |  | 195 (8.2) |
|  | Salary | 67 (15.2) | 333 (17.1) | <0.001 | 400 (16.8) |
| Full/Part-time status | | | | | |
| (n=443) | Full time | 258 (58.2) | 1291 (66.3) |  | 1,549 (64.8) |
|  | Part time | 125 (28.2) | 544 (27.9) |  | 669 (28.0) |
|  | Other | 60 (13.5) | 113 (5.8) | <0.001 | 173 (7.2) |
| Customer Status | | | | | |
| (n=459) | No customers | 105 (22.9) | 697 (23.9) |  | 802 (23.8) |
|  | Yes customers | 354 (77.1) | 2219 (76.1) | 0.631 | 2573 (76.2) |
| Organization Size | | | | | |
| (n=451) | 1 to 10 | 49 (10.9) | 391 (13.8) |  | 440 (13.4) |
|  | 11 to 49 | 157 (34.8) | 357 (12.6) |  | 1114 (34.0) |
|  | 50 - 499 | 211 (46.8) | 1237 (43.7) |  | 1448 (44.2) |
|  | >=500 | 34 (7.5) | 244 (8.6) | 0.26 | 278 (8.5) |
| USDA Food Security Score | | | | | |
| (n=414) | High or marginal | 234 (56.5) | 1065 (60.2) |  | 1299 (59.5) |
|  | Low food security | 84 (20.3) | 379 (21.4) |  | 463 (21.2) |
|  | Very low food security | 96 (23.2) | 325 (18.4) | 0.082 | 421 (19.3) |
| " If I was sick with COVID-19, but I was still able to work, I would go to work" | |  |  |  |  |
| (n=2,535) Agree  Do not agree | | 40 (9.0)  405 (91.0) | 182 (8.7)  1,908 (91.3) | 0.849 | 222 (8.8)  2,313 (91.2) |
